# Is psychomotor retardation associated with treatment response in adults with depression? A secondary analysis of nine randomised controlled trials

**DOI:** 10.64898/2026.01.07.25342551

**Authors:** James B Badenoch, Lucy I Stiles, Anthony S David, Glyn Lewis, Joshua EJ Buckman, Jonathan P Rogers

## Abstract

**Background:** Psychomotor retardation is characterised by slowed motor and cognitive processes. Although associated with greater illness severity in depression, its relationship with treatment response is unclear. We examined whether psychomotor retardation is associated with treatment outcomes in depression.

**Methods:** We conducted a secondary analysis of individual participant data from nine randomised controlled trials of pharmacological, psychological and exercise-based treatments for unipolar depression in primary care. Self-reported psychomotor retardation was assessed at baseline using the Revised Clinical Interview Schedule. The primary outcome was the standardised *z*-score of each trial’s validated self-report depression measure at 3–4 months post-randomisation; the secondary outcome was remission. Within-study regression models were adjusted for established prognostic factors, with secondary analyses additionally adjusting for baseline core depressive symptoms and depression severity. Effect estimates were pooled using random-effects meta-analysis.

**Results:** Among 4,290 participants, 2,754 (64.2%) reported psychomotor retardation. Psychomotor retardation was associated with greater depression severity at follow-up (pooled *z*-score = 0.184, 95% CI 0.059–0.309, p = 0.004, *I*² = 68.7%) and lower odds of remission (pooled OR = 0.737, 95% CI 0.577–0.941, p = 0.015, *I*² = 57.7%), adjusting for age, sex, ethnicity, marital status, employment status and allocated treatment. Findings were robust to adjustment for baseline core depressive symptoms but not after adjustment for full baseline severity.

**Conclusions:** Psychomotor retardation is associated with poorer treatment outcomes in depression, though this may reflect underlying severity. Recognition of psychomotor retardation may help identify patients at risk of poorer response and inform clinical management.

## Introduction

Psychomotor retardation is characterised by the slowing of motor and cognitive processes including disturbances in speech, facial expression, and fine and gross motor behaviour (Bennabi et al., 2013). Psychomotor retardation is a common feature of depression experienced by around 29% of patients (Wüthrich et al., 2024) and higher rates (up to 73%) are observed in depression with psychotic features (Janzing et al., 2020). The *Diagnostic and Statistical Manual of mental disorders* (DSM-5) includes psychomotor retardation as one of the nine core symptoms of major depressive disorder (MDD) (American Psychiatric Association, 2013). Similarly, marked slowing of thought and movement is listed as one of the ICD-11 diagnostic criteria (World Health & Organization (WHO), 2019). Emerging evidence suggests that psychomotor retardation has distinct neurobiological underpinnings. Neuroimaging studies have implicated dysfunction in cortico-striatal motor circuits and structural changes in the basal ganglia (Osborne et al., 2020). Dopaminergic dysfunction may also play a role, with reduced dopamine synthesis and altered receptor availability observed in individuals with psychomotor retardation (Bennabi et al., 2013).

Psychomotor retardation is closely associated with depression severity and chronicity, with self-reported psychomotor retardation linked with earlier age of onset, longer illness duration, greater frequency of episodes and more lifetime suicidal behaviours (Calugi et al., 2011). These findings have been validated in a larger cohort, where subjectively reported psychomotor retardation was predictive of greater overall depressive symptom severity and disability (Gómez-Cumplido et al., 2024). Similarly, anhedonia is associated with psychomotor retardation in unipolar depression (Lemke et al., 1999). Objective measures of slowed movement are negatively associated with both self-reported and clinician-rated depression severity, including sadness (r = –0.35, p = 0.03) and feelings of failure (r = –0.35, p = 0.03), from the Beck Depression Inventory – Second Edition (BDI-II) (Beck et al., 2011).

Psychomotor retardation is strongly associated with treatment outcomes in depression. Studies have shown that psychomotor retardation is linked to poorer response to pharmacological treatment. In depression with psychotic features, psychomotor retardation has been associated with lower remission rates (Flint et al., 2022; Janzing et al., 2020) and higher persistence of delusions (Janzing et al., 2020). Paradoxically, multiple studies have shown that patients with psychomotor retardation experience greater symptom reduction and higher response rates to ECT compared to those without psychomotor retardation (Heijnen et al., 2019; van Diermen et al., 2019).

Existing evidence links psychomotor retardation to greater depression severity and poorer treatment outcomes. However, it remains unclear whether psychomotor retardation is independently associated with poorer prognosis. Existing studies are often limited by small sample sizes. Clarifying whether psychomotor retardation is an independent prognostic factor is clinically important. If psychomotor retardation is reliably associated with poorer outcomes, it could help clinicians identify patients who may require earlier, tailored or augmented treatment regimens and more frequent monitoring, and may also support clearer communication with patients about the likely course of their recovery. To address this, our study aims to investigate whether self-reported psychomotor retardation is independently associated with treatment outcomes in depression, using individual participant-level data from nine randomised controlled trials investigating treatment for depression and adjusting for multiple established prognostic variables.

## Methods

This study is a secondary analysis of individual participant data from nine randomised controlled trials (RCTs) included in the Depression in General Practice (Dep-GP) individual participant data dataset, which originally included thirteen RCTs of depression treatments in primary care. The Dep-GP dataset has been used for Individual Participant Data Meta-Analysis in the past (Buckman, Saunders, Cohen, et al., 2021). The interventions consisted of pharmacological, psychological and physical exercise-based therapies. Inclusion of studies from the Buckman et al., 2021 Individual Participant Data Meta-Analysis (IPDMA) was based on the systematic use of the Revised Clinical Interview Schedule (CIS-R) at baseline and a lack of systematically missing outcome or covariate data. Based on this, four RCTs were not eligible for inclusion due to a lack of primary outcome measure at three-four months post-intervention (CPN-GP and ITAS) (Kendrick et al., 2005, Thomas et al., 2004), lack of an exposure measure for psychomotor retardation (AHEAD) (Kendrick et al., 2006) and systematically missing covariate data (Healthlines) (Salisbury et al., 2016). Individuals missing data for psychomotor retardation were excluded from the analysis.

The primary exposure was the presence or absence of subjective psychomotor retardation, assessed at baseline using the CIS-R psychomotor retardation question “When you have felt sad, miserable or depressed/unable to enjoy or take an interest in things in the past seven days, have you been doing things more slowly, for example, walking more slowly?”. Responses were coded as a binary variable (present or absent). The primary outcome was a continuous variable (the standardised z-score of the primary outcome measure used in each RCT). The secondary outcome, remission, was defined as a binary variable (remission versus no remission) based on trial-specific thresholds (as per Buckman et al., 2021). Covariates included age (continuous), self-identified sex (binary – male or female), ethnicity (binary – white or non-white), marital status (categorical – married/co-habitating or single or divorced), employment status (categorical – employed or not seeking employment or unemployed), and treatment allocation (randomised treatment modality); categorisations were consistent with previous studies in this dataset (Buckman et al., 2021).

Individuals with psychomotor agitation were coded as not having psychomotor retardation, while individuals with both psychomotor agitation and retardation were excluded. The primary outcome was the standardised z-score of the primary outcome measure used by individual RCTs at three to four months post-randomisation, to harmonise assessment measures across studies. Secondary outcome measures included remission as defined by the individual RCTs (binary variable) and a Global Rating of Change (GRC) – the latter of which was dichotomised into ‘feeling better’ and ‘not feeling better’ based on (Buckman et al., 2022). GRC data was only available for two RCTs: PANDA (Lewis et al., 2019) and MIR (D. S. Kessler et al., 2018).

### Statistical analysis

For the primary analysis, regression models were fitted within studies to evaluate the effect of psychomotor retardation on the standardised z-score of the primary outcome measure (linear model) and remission (logistic model). Models were adjusted for age, sex, ethnicity, employment status, marital status and specific randomised treatment; this was based on known prognostic factors in unipolar depression, validated in this cohort (Buckman, Saunders, Stott, et al., 2021). To adjust for the effect of depression severity, secondary analyses included baseline core depressive symptoms (as measured by the CIS-R depression score – including the anhedonia and low mood items) as covariates. A further model included the standardised z-score of the baseline depression severity measure as a covariate (consistent with the outcome measure used).

Two studies (CADET and REACT) assessed baseline depression severity using the PHQ-9, which includes an item that measures psychomotor symptoms (“moving or speaking slowly, or being fidgety or restless”). Because this item captures both psychomotor retardation and agitation, adjusting for the total PHQ-9 score at baseline risked introducing collinearity and partial adjustment for our exposure of interest. To address this, we recalculated baseline and follow-up PHQ-9 total scores with the psychomotor item removed and used these revised totals to generate standardised depression severity scores for use as covariates in the sensitivity analyses.

To test the effect of psychomotor retardation on Global Rating of Change – the secondary outcome measure – a logistic regression model was used and adjusted for age, sex, ethnicity, employment status, marital status and specific randomised treatment.

Meta-analysis was conducted to pool the within-study regression estimates from the linear and logistic models, adjusted for core depressive symptoms at baseline. A sensitivity meta-analysis used pooled data, which were adjusted for baseline depression severity (z-score of the baseline depression severity measure). A random-effects model was used, and between-study variance was evaluated using the restricted maximum likelihood estimator method (Cooper et al., 2009). The I^2^ statistic was used to quantify the proportion of variation in effect sizes that is likely due to between-study variability. Forest plots with 95% confidence intervals were produced for each meta-analysis.

In terms of missing data, RCTs with systematically missing relevant data (Healthlines and ITAS) were not included in this study. Multivariate imputation by chained equations (MICE) was used for non-systematically missing variables with five imputations, at which point the estimates converged. The imputed dataset was used in a sensitivity meta-analysis to compare whether missing data impacted the observed associations in the model adjusted for baseline core depressive symptoms. Effect sizes were reported as regression coefficients or odds ratios with 95% confidence intervals. The threshold for statistical significance was set to p < 0.05. Analysis was conducted in R version 2025.5.1.513 (R Core Team, 2024). The MICE package version 3.16.0 was used for multiple imputation (van Buuren, 2024). The metafor package version 4.8-0 was used for meta-analysis (Viechtbauer, 2010).

## Results

### Descriptive Statistics

Four RCTs included in the original IPDMA were excluded from this study. Two were not eligible for inclusion due to a lack of primary outcome measure at three-four months post-intervention (ITAS, n = 798; CPN-GP, n = 247) (Thomas et al., 2004). Another had a lack of covariate data (Healthlines, n = 609) (Salisbury et al., 2016), and a final study was excluded as it did not report item-level CIS-R data required to derive a psychomotor retardation variable (AHEAD, n = 327) (Kendrick et al., 2006). Individuals with both psychomotor agitation and retardation were excluded (n = 281, across all eligible studies). The final study population included 4,290 participants, of whom 2,754 (64%) had psychomotor retardation – with some variation in the frequency of psychomotor retardation between studies (Supplementary Table 2). The baseline characteristics of the study population are summarised in Table 2. The number of participants with missing data for each variable is included in Supplementary Table 1.

**Table 1.** Included Studies.

|  |  |  | Inclusion criteria | Age | Sex | Baseline Depression symptom severity | Remission |  | Outcome measure | Included (Y/N) | Reason for exclusion |
| --- | --- | --- | --- | --- | --- | --- | --- | --- | --- | --- | --- |
| Study | N | Pragmatic RCT (Y/N) |  | Mean (S.D.) | % Female | Mean (S.D.) | % at 3-4 months | Interventions | Primary (additional) |  |  |
| AHEAD (Kendrick et al., 2006) | 327 | Y | Adults with new depressive episodes diagnosed by GP | 43.1 (15.4) | 67 | HADS 10.5 (3.9) | 62 | TCA vs. SSRI vs. Lofepramine | HADS (CIS-R) | N | Did not report item-level CIS-R data required to derive a psychomotor retardation variable |
| CADET (Richards et al., 2013) | 527 | Y | Adults ≥18, ICD-10 Depressive episodes | 44.4 (13.2) | 72 | PHQ-9 17.7 (5.1) | 41 | Collaborative care v. TAU | PHQ-9 | Y | - |
| COBALT (Wiles et al., 2013) | 469 | Y | Adults 18-75 with treatment resistant depression, scoring ≥14 BDI-II | 49.6 (11.7) | 72 | BDI-II 31.8 (10.7) | 34 | CBT + TAU v. TAU | BDI-II (PHQ-9) | Y | - |
| CPN-GP (Kendrick et al., 2006) | 247 | Y | Adults 18-65 seeking treatment for new episode of depression, anxiety or reaction to life difficulties, scoring >3 on GHQ-12 | - | 67 | GHQ-12 | - | TAU vs CPN vs CPN + Problem solving | CIS-R (and HADS) | N | No primary outcome measure at 3-4 months |
| GENPOD (Wiles et al., 2012) | 601 | N | Adults 18-74 with depressive episode | 38.8 (12.4) | 68 | BDI-II 33.7 (9.7) | 41 | Citalopram v. reboxetine | BDI-II (HADS) | Y | - |
| HEALTHLINES (Salisbury et al., 2016) | 609 | Y | Adults ≥18, PHQ-9 ≥10, confirmed diagnosis of depression with CIS-R, internet access | 49.5 (12.9) | 69 | PHQ-9 16.9 (4.6) | 30 | Healthlines telecare + TAU v. TAU | PHQ-9 | N | Systematically missing covariate data |
| IPCRESS (D. Kessler et al., 2009) | 295 | Y | Adults scoring ≥14 BDI-II and GP confirmed diagnosis of depression | 34.9 (11.6) | 68 | BDI-II 33.2 (8.8) | 34 | iCBT + TAU v. TAU + waiting list for iCBT | BDI-II | Y | - |
| ITAS (Thomas et al., 2004) | 798 | Y | Adults $\geq 16$ scoring $\geq 12$ on CIS=R | 43.2 (14.8) | 68 | GHQ 7.7 (3.2) | N/A; at 6-8 months: 46 | Recommendation + TAU v. TAU | GHQ-12 | N | No primary outcome measure at 3-4 months |
| MIR (D. S. Kessler et al., 2018) | 480 | Y | Adults $\geq 18$ taking SSRIs or SNRIs at adequate dose for $\geq 6$ weeks and scored $\geq 14$ on BDI-II | 50.7 (13.2) | 69 | BDI-II 31.1 (9.9) | 30 | Mirtazapine v. placebo | BDI-II (PHQ-9) | Y | - |
| PANDA (Lewis et al., 2019) | 652 | Y | Adults presenting with low mood or depression to GP in last 3 years, free of ADM for 8 weeks up to baseline | 39.7 (15.0) | 59 | BDI-II 23.9 (10.3) | 69 | Sertraline v. placebo | PHQ-9 (BDI-II) | Y | - |
| REEACT (Gilbody et al., 2015) | 685 | Y | Adults with PHQ-9 $\geq 10$ presenting to GP with depression | 39.9 (12.7) | 67 | PHQ-9 16.7 (4.3) | 53 | Moodgym v. beating the blues v. TAU | PHQ-9 | Y | - |
| RESPOND (Sharp et al., 2010) | 220 | Y | Women meeting criteria for MDD within 6-months post-partum | 28.7 (6.4) | 100 | EPDS 17.6 (3.4) | 56 | ADM v. listening intervention | EPDS | Y | - |
| TREAD (Chalder et al., 2012) | 361 | Y | Adults 18-69 who met diagnostic criteria for MDD and scored $\geq 14$ on BDI-II | 39.8 (12.6) | 66 | BDI-II 32.1 (9.2) | 35 | Physical activity + TAU v. TAU | BDI-II | Y | - |

ADM, Antidepressant medication; BDI-II, Beck Depression Inventory-II; CBT, Cognitive behavioural therapy; CIS-R, Revised Clinical Interview Schedule; EPDS, Edinburgh Postnatal Depression Scale; GHQ-12, General Health Questionnaire (12 items); GP, General practitioner; HADS, Hospital Anxiety and Depression Scale; HADS-D, Hospital Anxiety and Depression Scale – Depression subscale; ICD-10, International Classification of Diseases, 10th Edition; iCBT, Internet-based therapist-delivered cognitive behavioural therapy; MDD, Major Depressive Disorder; PHQ-9, Patient Health Questionnaire-9; RCT, Randomised controlled trial; SSRI, Selective serotonin reuptake inhibitor; SNRI, Serotonin-norepinephrine reuptake inhibitor; TAU, Treatment as usual; TCA, Tricyclic antidepressant..

**Table 2.** Baseline Characteristics.

| **Variable** | **Psychomotor retardation**<br>**absent**<br>**(n = 1536)** | **Psychomotor retardation**<br>**present**<br>**(n = 2754)** |
| --- | --- | --- |
| Age, mean (SD) | 40.1 (13.2) | 42.5 (14.1) |
| Sex, n (%) |  |  |
| - Female | 1093 (71) | 1866 (68) |
| - Male | 443 (29) | 887 (32) |
| Ethnicity, n (%) |  |  |
| - White | 1434 (93) | 2560 (93) |
| - Non-white | 100 (7) | 194 (7) |
| Employment status, n (%) |  |  |
| - Employed | 924 (60) | 1375 (50) |
| - Not seeking<br>employment | 389 (25) | 774 (28) |
| - Unemployed | 222 (15) | 602 (22) |
| - Marital status, n (%) |  |  |
| - Married/co-habit | 756 (49) | 1344 (49) |
| - Single | 512 (34) | 829 (35) |
| - Divorced | 266 (17) | 579 (17) |
| Treatment allocation, n (%) |  |  |
| - TAU | 535 (35) | 870 (32) |
| - Antidepressants | 479 (31) | 1034 (38) |
| - Psychological<br>therapy | 365 (30) | 725 (26) |
| - Other | 57 (3) | 125 (5) |
| CIS-R depression score, mean (SD) |  |  |
| CIS-R depression score (anhedonia and low mood only) | 2.45 (1.29) | 3.08 (0.95) |
| Baseline depression severity*, mean (SD) | -0.27 (0.94) | 0.25 (0.94) |

\*Z-score of primary measure

### Association between psychomotor retardation and depression severity or remission at follow-up

Psychomotor retardation at baseline showed statistical evidence of an association with increased depression severity at follow-up in four studies including CADET (z-score = 0.27, 95% CI [0.07, 0.47]), PANDA (z-score = 0.43, 95% CI [0.28, 0.58]), REEACT (z-score = 0.34, 95% CI [0.19, 0.50]), and RESPOND (z-score = 0.32, 95% CI [0.01, 0.63]) (Table 3). A random-effects meta-analysis showed a significant pooled effect (z-score = 0.18, 95% CI [0.06, 0.31]), with moderate heterogeneity (I² = 68.7%, 95% CI [30.3%, 92.5%]) (Figure 1). These models were adjusted for age, sex, ethnicity, employment, marital status and treatment allocation.

**Figure 1:**
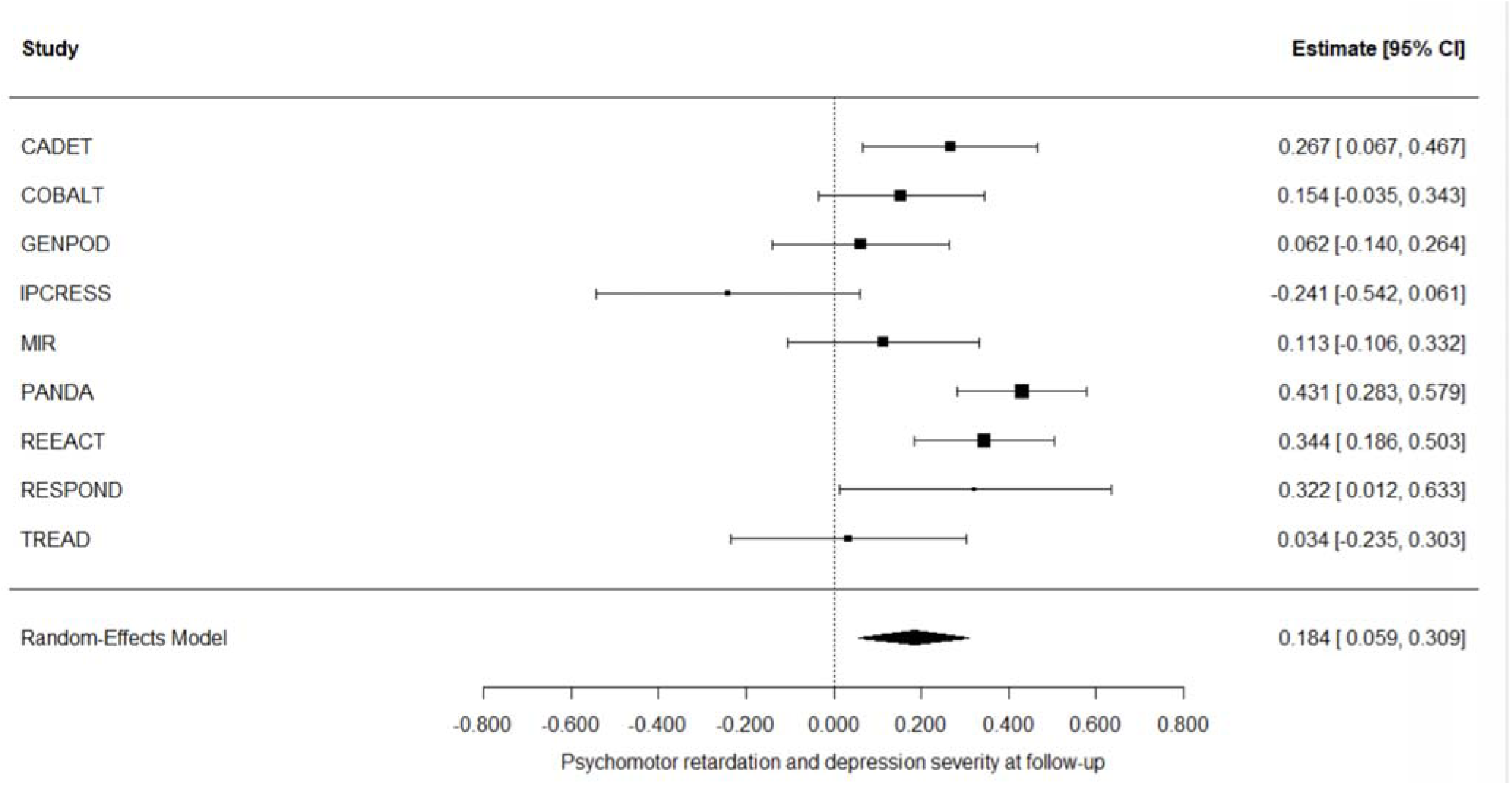
forest plot psychomotor retardation and primary outcome measure. p = 0.004, I^2^ 68.7% (I^2^ 95% CI 30.3 – 92.5%)

**Table 3.** Within-study adjusted mean differences for psychomotor retardation and depression severity at follow-up.

| Study | Estimate | 95% CI (Low) | 95% CI (High) | P-value | n |
| --- | --- | --- | --- | --- | --- |
| **CADET** | 0.27 | 0.07 | 0.47 | < 0.01 | 527 |
| **COBALT** | 0.15 | -0.04 | 0.34 | 0.11 | 469 |
| **GENPOD** | 0.06 | -0.14 | 0.26 | 0.55 | 601 |
| **IPCRESS** | -0.24 | -0.54 | 0.06 | 0.12 | 295 |
| **MIR** | 0.11 | -0.11 | 0.33 | 0.31 | 480 |
| **PANDA** | 0.43 | 0.28 | 0.58 | < 0.001 | 652 |
| **REEACT** | 0.34 | 0.19 | 0.50 | < 0.001 | 685 |
| **RESPOND** | 0.32 | 0.01 | 0.64 | < 0.05 | 220 |
| **TREAD** | 0.03 | -0.24 | 0.30 | 0.80 | 361 |

Psychomotor retardation at baseline was similarly associated with a lower odds of remission in three studies including CADET (OR = 0.582, 95% CI [0.385, 0.880]), PANDA (OR = 0.386, 95% CI [0.259, 0.570]), and REEACT (OR = 0.599, 95% CI [0.416, 0.860]) (Table 4). The pooled odds ratio from the random-effects model was 0.737 (95% CI [0.577, 0.941], p < 0.05), indicating a significant association between psychomotor retardation and reduced likelihood of remission, with moderate heterogeneity (I² = 57.7%, 95% CI [6.66%, 87.8%]) (Figure 2). These models were adjusted for age, sex, ethnicity, employment, marital status and treatment allocation.

**Figure 2:**
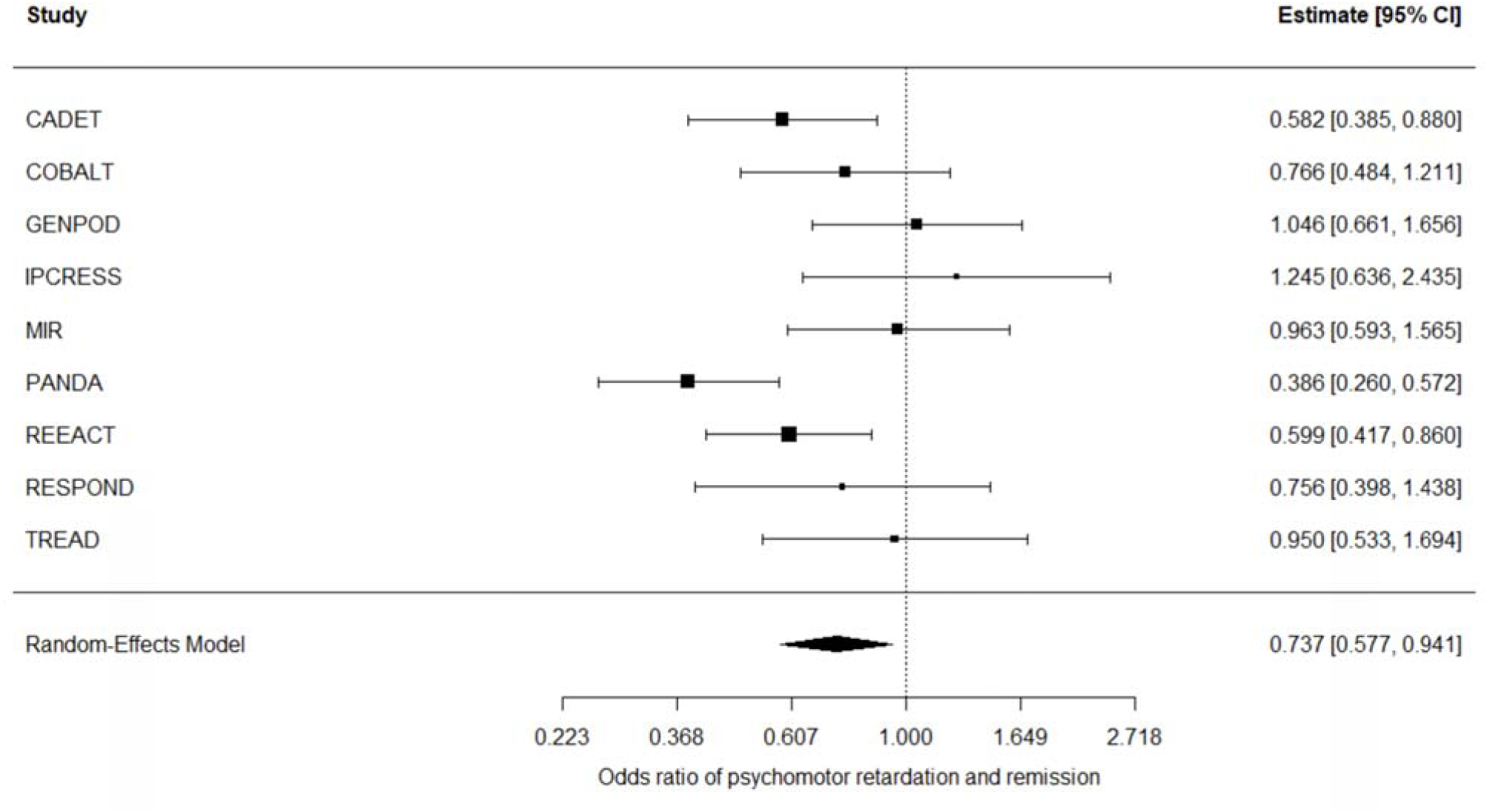
forest plot psychomotor retardation and remission. p = 0.015, I^2^ 57.7% (I^2^ 95% CI 6.66 – 87.8%)

**Table 4.**
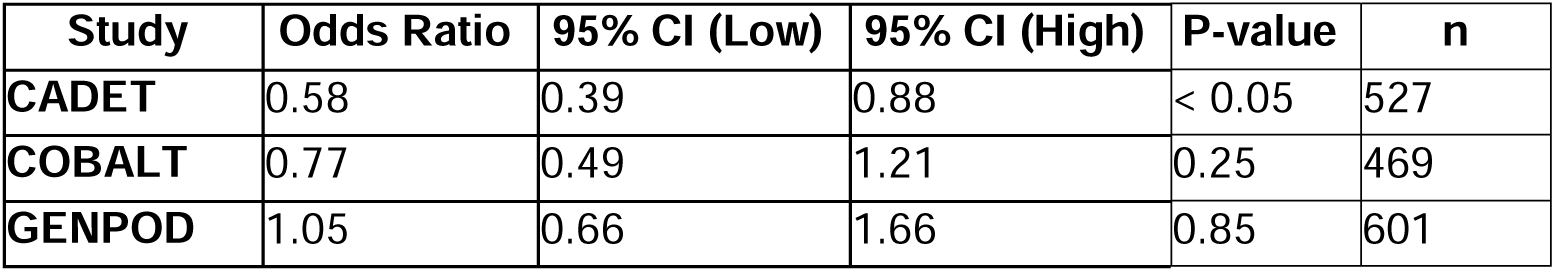
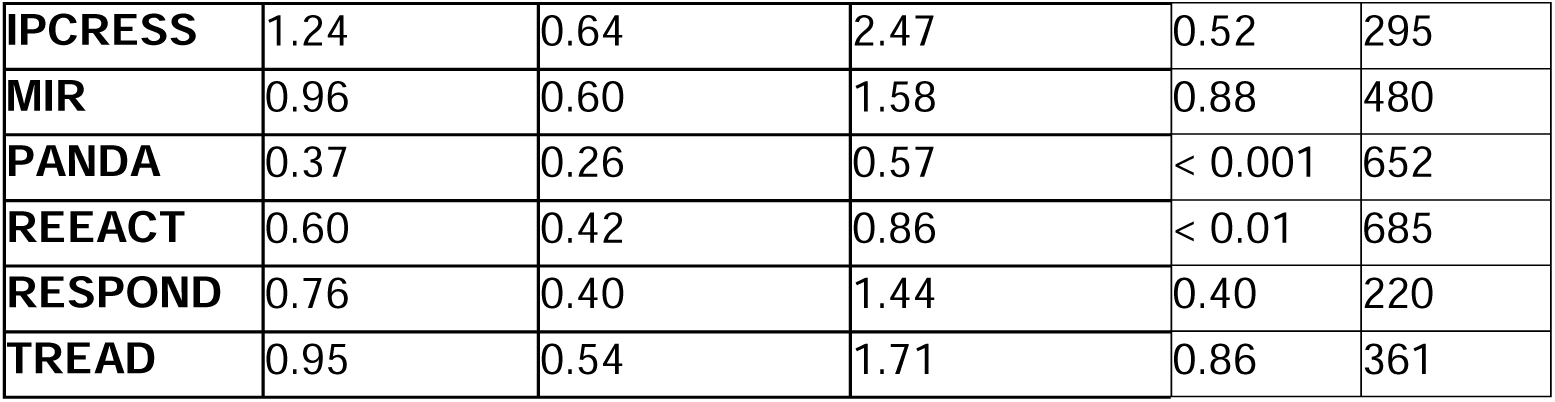
Within-study odds ratios for psychomotor retardation and remission at follow-up.

### Adjusting for core depressive symptoms

When additionally adjusting for baseline core depressive symptoms (CIS-R depression score), the association between psychomotor retardation and depression severity at follow-up remained significant in studies including CADET (z-score = 0.23, 95% CI [0.04, 0.43]), PANDA (z-score = 0.19, 95% CI [0.04, 0.35]), and REEACT (z-score = 0.27, 95% CI [0.10, 0.44]). However, a negative association was observed in IPCRESS (β = –0.31, 95% CI [–0.61, –0.01]) (Table 5). The adjusted random-effects meta-analysis remained significant (z-score = 0.12, 95% CI [0.03, 0.21], p < 0.01), with moderate heterogeneity (I² = 39.5%, 95% CI [0%, 89.2%]) (Figure 3).

**Figure 3:**
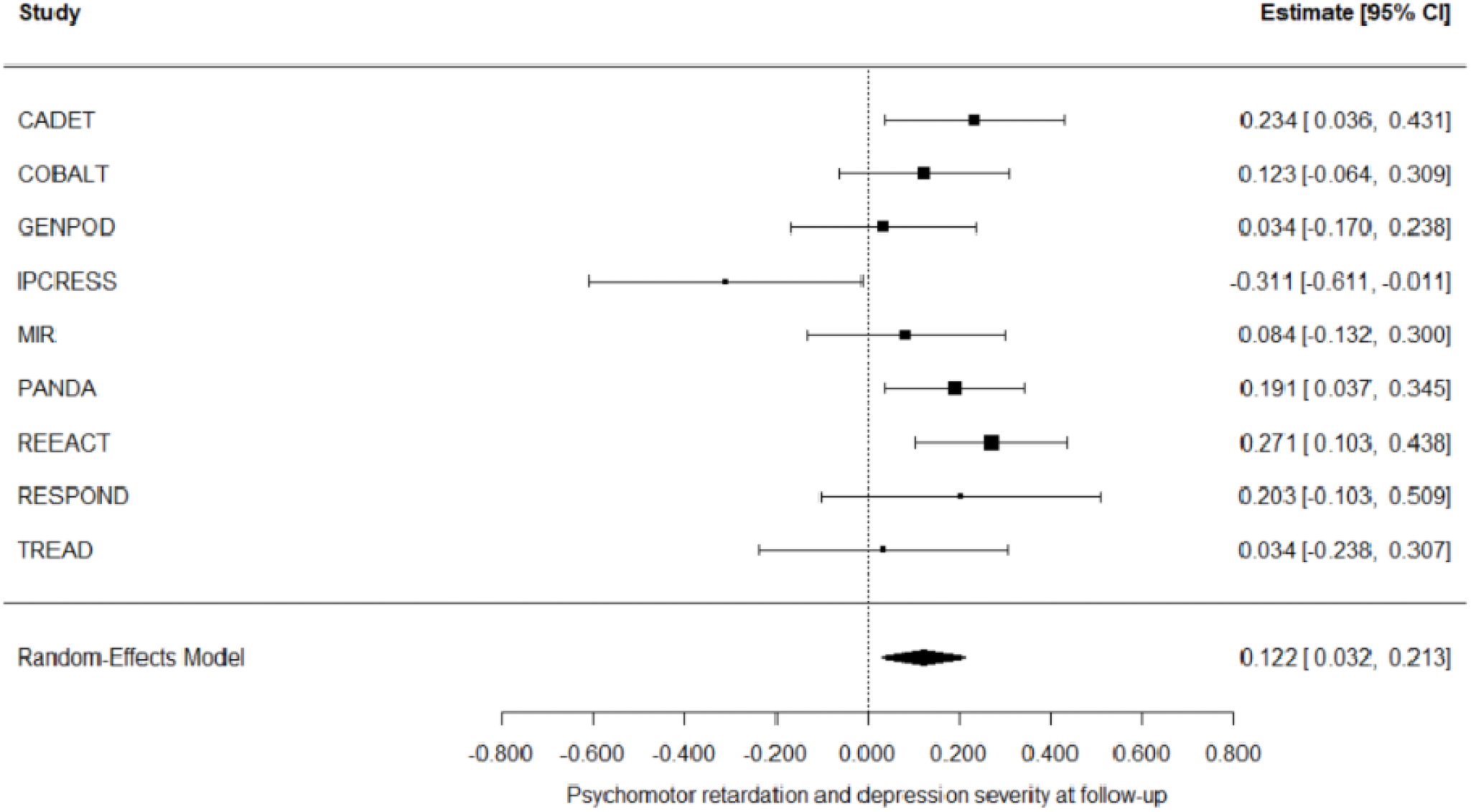
forest plot psychomotor retardation and primary outcome measure (adjusting for CIS-R depression score) p < 0.01, I^2^ 39.5% (I^2^ 95% CI 0 – 89.2%)

**Table 5.**
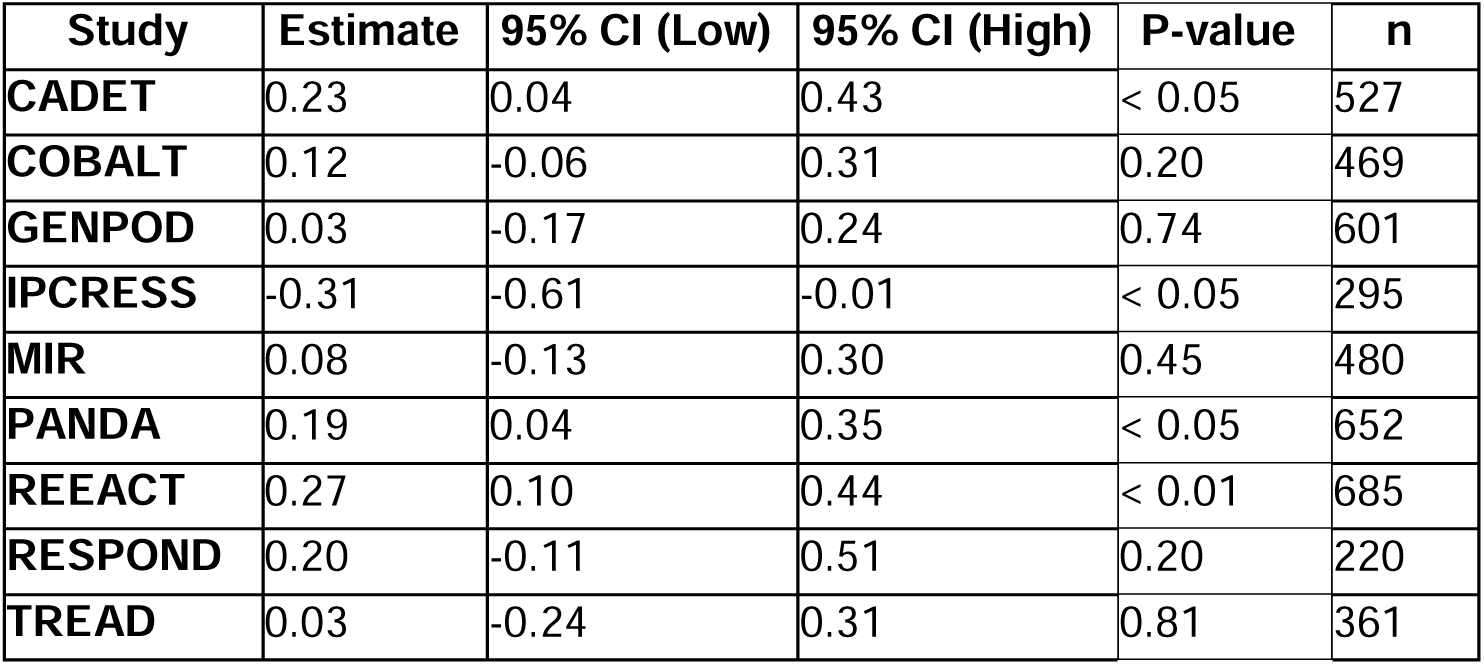
Within-study adjusted mean differences for psychomotor retardation and Depression Severity at Follow-up (adjusting for baseline CIS-R score)

This was similarly found with remission as the outcome measure: CADET (OR = 0.58, 95% CI [0.39, 0.88]), PANDA (OR = 0.39, 95% CI [0.26, 0.57]), and REEACT (OR = 0.60, 95% CI [0.42, 0.86]) (Table 6). The pooled odds ratio remained significant (OR = 0.81 95% CI [0.68, 0.97], p < 0.05) with low heterogeneity (I² = 19.3%, 95% CI [0%, 77.2%]) (Figure 4).

**Figure 4:**
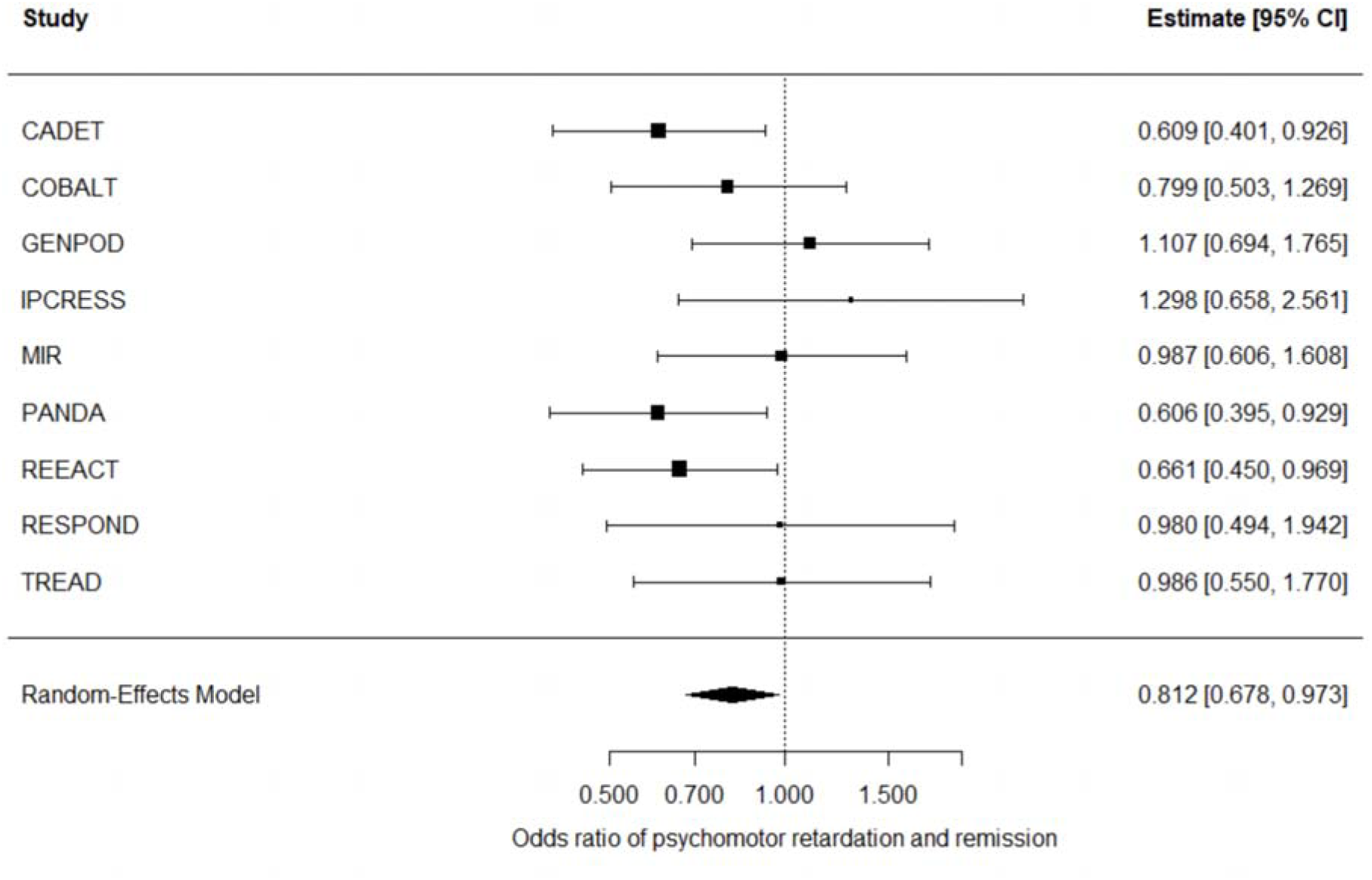
forest plot psychomotor retardation and remission (adjusting for CIS-R depression score) p < 0.05, I^2^ 19.3% (I^2^ 95% CI 0 – 77.2%)

**Table 6.**
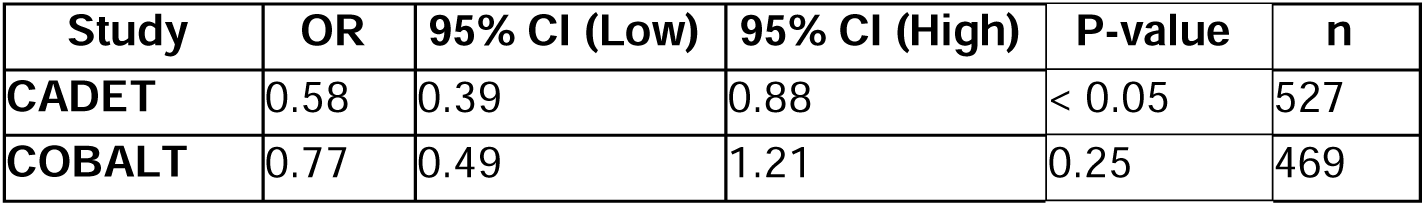
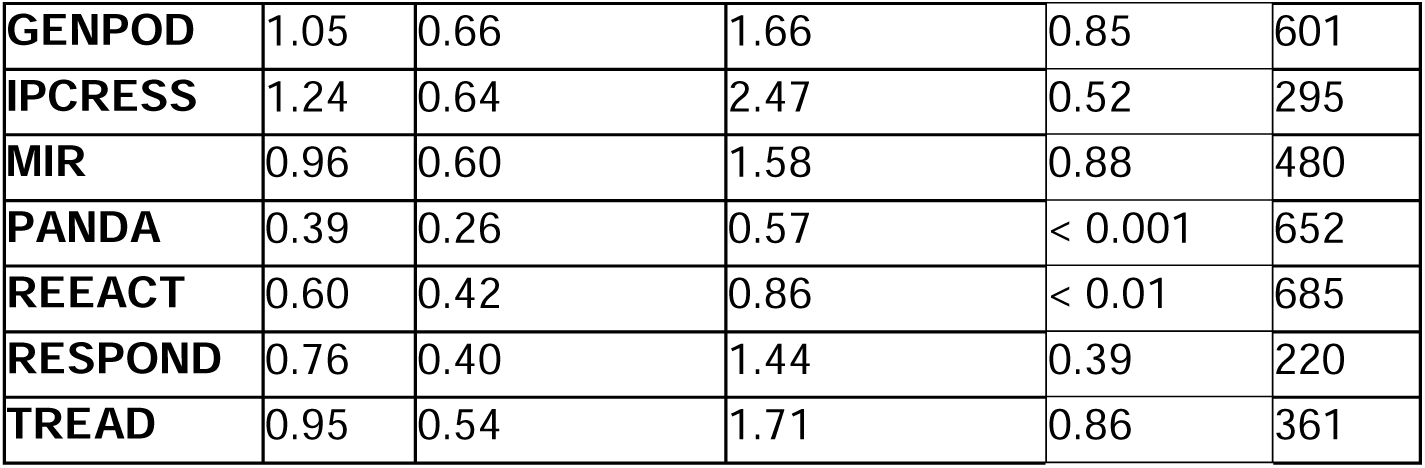
Within-study adjusted mean differences for Psychomotor Retardation and Remission (adjusting for baseline CIS-R score)

| **Study** | **OR** | **95% CI (Low)** | **95% CI (High)** | **P-value** | **n** |
| --- | --- | --- | --- | --- | --- |
| **CADET** | 0.58 | 0.39 | 0.88 | < 0.05 | 527 |
| **COBALT** | 0.77 | 0.49 | 1.21 | 0.25 | 469 |
| **GENPOD** | 1.05 | 0.66 | 1.66 | 0.85 | 601 |
| **IPCRESS** | 1.24 | 0.64 | 2.47 | 0.52 | 295 |
| **MIR** | 0.96 | 0.60 | 1.58 | 0.88 | 480 |
| **PANDA** | 0.39 | 0.26 | 0.57 | < 0.001 | 652 |
| **REEACT** | 0.60 | 0.42 | 0.86 | < 0.01 | 685 |
| **RESPOND** | 0.76 | 0.40 | 1.44 | 0.39 | 220 |
| **TREAD** | 0.95 | 0.54 | 1.71 | 0.86 | 361 |

### Adjusting for baseline depression severity

A sensitivity analysis additionally adjusted for baseline depression severity (mean z-score of the individual study baseline depression severity measure). The within-study models were broadly consistent with the CIS-R adjusted results (Supplementary Table 2, Supplementary Table 3). In the pooled results, psychomotor retardation associations lost significance (z-score 0.03, 95% CI [-0.08, 0.14], p = 0.56, I² = 61.8%) and remission (OR 0.91, 95% CI [0.76, 1.10], p = 0.34, I² = 19.5%) (Supplementary Figure 1, Supplementary Figure 2).

### Sensitivity analysis using imputed data

A sensitivity analysis using a dataset with five imputations demonstrated consistent results with the main analysis; psychomotor retardation remained significantly associated with greater depression severity at follow-up (pooled z-score = 0.12, 95% CI [0.03, 0.20], p < 0.01, I² = 46.1%) and remission (pooled OR = 0.82, 95% CI [0.70, 0.95], p < 0.05, I² = 9.78%), when adjusting for baseline core depressive symptoms (CIS-R score) (Figure S3, S4).

### Secondary outcome (global rating of change)

There was no association between psychomotor retardation and Global Rating of Change (“feeling better”) in either the PANDA (OR 0.77, 95% CI [0.54 - 1.09], p = 0.145) or MIR (OR 1.37, 95% CI [0.83 - 2.24], p = 0.21) studies, adjusting for age, ethnicity, employment status, allocated treatment, sex and marital status. The pooled odds ratio from the random effects meta-analysis 0.998 (95% CI [0.57 - 1.75), p = 0.995, I² =71.1% (95% CI [0%, 99%]).

## Discussion

This study examined the role of psychomotor retardation as a prognostic indicator in adults with unipolar depression by analysing individual participant level data from nine RCTs. The presence of psychomotor retardation at baseline was associated with more severe symptoms and lower odds of remission at follow-up, after adjusting for known prognostic factors of depression, treatment allocation and baseline depressive symptoms. Overall, the results support psychomotor retardation as a meaningful indicator of poorer prognosis, independent of modality of treatment. The magnitude of the association was attenuated after adjusting for baseline depression severity and there was no identified association between baseline psychomotor retardation and a patient-rated measure of improvement. Psychomotor retardation may not therefore add substantial predictive power to overall severity.

### Neurobiological underpinnings of psychomotor retardation in depression

Emerging evidence implicates structural and functional brain changes, along with predominately dopaminergic dysfunction in psychomotor retardation. Structural and functional abnormalities across cortico-striatal-thalamo-cortical (CSTC) motor circuits, particularly involving the striatum, caudate, putamen, and supplementary motor area (SMA), disrupt the integration of motor planning and execution of voluntary movement. Evidence highlights a specific role for dopaminergic dysregulation in psychomotor retardation including reduced dopamine synthesis in the caudate (Martinot et al., 2001) and increased D2 receptor availability in the striatum (Shah et al., 1997). Similarly, in the putamen there is increased [11C]-raclopride binding in psychomotor retardation (Meyer et al., 2006) and structural volume loss in MDD (Husain et al., 1991).

### Treatment stratification

Distinct biological mechanisms underlying psychomotor retardation from other depressive symptoms may account for the poorer outcomes observed in patients with psychomotor retardation, though it is difficult to study this where there are few head-to-head comparisons of antidepressants with different mechanisms. Five of the included RCTs evaluated the efficacy of antidepressant treatment, and these were predominantly serotonergic and noradrenergic modulators (SSRIs, TCAs and SNRIs). Minimal dopaminergic modulation is a possible explanation for the poorer treatment response amongst individuals with psychomotor retardation.

This has important implications for treatment stratification; individuals with prominent psychomotor retardation may require neurobiologically informed interventions that directly target motor circuitry and dopaminergic function. There is some evidence that imipramine – which has a modest inhibitory effect on dopaminergic synaptic reupdate (Stille & Michaelis, 1970) - was associated with lower levels of psychomotor retardation compared to individuals taking venlafaxine (Janzing et al., 2020). Bupropion, a noradrenaline-dopamine reuptake inhibitor, could confer therapeutic effects in this patient population given its direct enhancement of dopaminergic transmission coupled with its established antidepressant effects (Patel et al., 2016). Dopamine agonists, such as bromocriptine, offer a mechanistically plausible approach to treating psychomotor retardation, as they directly stimulate post-synaptic dopamine receptors and exert antidepressant effects (Nordin et al., 1981). Dopamine agonists are well-established in the treatment of motor symptoms in conditions such as Parkinson’s disease (Tanner, 2000). Given the mechanistic overlap of psychomotor retardation and Parkinson’s disease (Leong et al., 2025), similar pharmacological approaches could hold therapeutic value for individuals with depression presenting with prominent psychomotor retardation. A large recent RCT of the dopamine agonist pramipexole supported its use in treatment-resistant depression, although it is currently unclear what the predictors of response are (Browning et al., 2025).

Aside from psychopharmacological interventions, electroconvulsive therapy is a particularly effective treatment for severe depression in those with psychomotor retardation (Heijnen et al., 2019). Electroconvulsive therapy enhances functional connectivity in frontal brain networks (Chen et al., 2023) and increases dopamine neurotransmission in the striatum (Landau et al., 2011), which could plausibly explain its particularly beneficial effect on depressed individuals with psychomotor retardation.

### Other Clinical and Theoretical Implications

Psychomotor retardation in depression is associated with distinct structural and functional brain changes, including frontostriatal abnormalities and impaired dopaminergic transmission (Bennabi et al., 2013). The presence of psychomotor retardation in depression has been associated with poorer treatment outcomes, including a delayed response to treatment with either interpersonal psychotherapy or selective serotonin reuptake inhibitor (SSRI) pharmacotherapy (Frank et al., 2011). Given the postulated neurobiological underpinnings of psychomotor retardation, it may represent a biologically distinct depression mechanism. Rather than being a distinct subtype, it is possible that patients lie along a mechanistic continuum, explaining some of the heterogeneity of the clinical presentation of depression as seen in people with psychomotor retardation including greater severity of depressive symptoms and divergence from typical treatment response patterns.

Currently, commonly used depression screening tools provide limited assessment of psychomotor retardation. The PHQ-9 and BDI-II contain one psychomotor retardation-related question, and the HADS omits psychomotor retardation entirely; however, the PHQ-9 combines psychomotor retardation with psychomotor agitation in a single question, reducing its specificity. Given our findings that psychomotor retardation is a significant predictor of treatment response, it is critical to identify at-risk patients. There is significant scope to revise screening tools to better identify psychomotor retardation, incorporating dedicated items or subscales.

### Strengths and Limitations

Individual participant-level data from nine RCTs allowed consistent adjustment for key covariates. However, as psychomotor retardation was not the randomised exposure, these analyses function observationally and remain susceptible to confounding. The inclusion of a range of treatment modalities, such as pharmacological, psychological and exercise interventions, enhances the generalisability of the findings across different therapeutic approaches. Psychomotor retardation was defined based on a self-reported measure using items of the CIS-R, this is likely to capture more subtle psychomotor symptoms which could be missed by a clinician-rate scale. However, this self-report measure of psychomotor retardation has questionable specificity given the high prevalence of psychomotor retardation (65%) in this cohort. It is unclear whether the results from a primary care population with what is likely to be mild psychomotor retardation (which was reported by patients themselves) generalises to more severe psychomotor (reported by clinicians) in secondary care. Additionally, a binary measure of psychomotor retardation does not capture severity or gradations of slowness. Several sensitivity analyses were used to interrogate the robustness of the association between psychomotor retardation and depression severity at follow-up.

## Conclusions and Future Directions

Psychomotor retardation is a significant predictor of poorer treatment outcomes and lower remission rates in individuals with unipolar depression, independent of core depressive symptoms and treatment modality. Neurobiological evidence implicating dopaminergic and motor-circuit dysfunction suggests that psychomotor retardation may represent a biologically distinct depression mechanism. Rather than being a distinct subtype, it is possible that patients lie along a mechanistic continuum, explaining some of the heterogeneity of the clinical presentation of depression. This may account for poorer outcomes with standard treatments which largely fail to target these mechanisms and encourages the use of targeted treatment options. Mechanistic studies are needed to further characterise dopaminergic dysfunction associated with psychomotor retardation, ideally in longitudinal cohorts, to better understand the temporal dynamics of psychomotor retardation and treatment response. Widely used depression screening tools lack sensitivity for early detection of individuals with a poorer prognostic trajectory and enable appropriate targeted treatment stratification.

## Data Availability

All data produced in the present study are available upon reasonable request to the authors

## Funding

JBB: NIHR ACF-2024-17-014.

JPR: NIHR CL-2023-18-002; Wellcome 220659/Z/20/Z

JEJB: Royal College of Psychiatrists, Medical Research Council, NIHR.

GL: NIHR, Wellcome Trust, UCLH BRC

