## Supplementary materials for "Is psychomotor retardation associated with treatment response in adults with depression? A secondary analysis of nine randomised controlled trials"

**Table S1 – Number and proportion of participants in each study with missing data by variable**

| **Study** | **Ethnicity n (%)** | **Psychomotor retardation at 3–4 months (log) n (%)** | **Depression severity at 6–8 months (z) n (%)** | **Employment status n (%)** | **Marital status n (%)** | **Sex n (%)** |
| --- | --- | --- | --- | --- | --- | --- |
| **CADET** | 0 (0.00%) | 37 (7.02%) | 527 (100.00%) | 2 (0.38%) | 0 (0.00%) | 0 (0.00%) |
| **COBALT** | 0 (0.00%) | 29 (6.18%) | 50 (10.66%) | 0 (0.00%) | 0 (0.00%) | 0 (0.00%) |
| **GENPOD** | 0 (0.00%) | 115 (19.13%) | 601 (100.00%) | 0 (0.00%) | 0 (0.00%) | 0 (0.00%) |
| **HEALTHLINES** | 2 (0.33%) | 86 (14.38%) | 93 (15.55%) | 7 (1.17%) | 598 (100.00%) | 0 (0.00%) |
| **IP CRESS** | 0 (0.00%) | 89 (30.17%) | 92 (31.19%) | 0 (0.00%) | 0 (0.00%) | 0 (0.00%) |
| **ITAS** | 798 (100.00%) | 798 (100.00%) | 210 (26.32%) | 1 (0.13%) | 3 (0.38%) | 0 (0.00%) |
| **MIR** | 1 (0.21%) | 56 (11.67%) | 88 (18.33%) | 2 (0.42%) | 0 (0.00%) | 0 (0.00%) |
| **PANDA** | 0 (0.00%) | 128 (19.63%) | 652 (100.00%) | 0 (0.00%) | 0 (0.00%) | 0 (0.00%) |
| **REEACT** | 0 (0.00%) | 163 (23.80%) | 685 (100.00%) | 0 (0.00%) | 1 (0.15%) | 0 (0.00%) |
| **RESPOND** | 1 (0.45%) | 35 (15.91%) | 220 (100.00%) | 0 (0.00%) | 3 (1.36%) | 0 (0.00%) |
| **TREAD** | 0 (0.00%) | 73 (20.22%) | 139 (38.50%) | 0 (0.00%) | 0 (0.00%) | 2 (0.55%) |

**Table S2: Frequency of Psychomotor Retardation by Study**

| **Study** | **Psychomotor retardation absent** | **Psychomotor retardation present** | **Total** |
| --- | --- | --- | --- |
| CADET, n (%) | 156 (29.6) | 371 (70.4) | 527 |
| COBALT, n (%) | 133 (28.4) | 336 (71.6) | 469 |
| GENPOD, n (%) | 126 (21.0) | 475 (79.0) | 601 |
| IPCRESS, n (%) | 96 (32.5) | 199 (67.5) | 295 |
| MIR, n (%) | 135 (28.1) | 345 (71.9) | 480 |
| PANDA, n (%) | 356 (54.6) | 296 (45.4) | 652 |
| REEACT, n (%) | 362 (52.8) | 323 (47.2) | 685 |
| RESPOND, n (%) | 80 (36.4) | 140 (63.6) | 220 |
| TREAD, n (%) | 92 (25.5) | 269 (74.5) | 361 |
| **Total, n (%)** | **1536 (35.8)** | **2754 (64.2)** | **4290** |

### **Table S3: Study-specific Regression Estimates for Psychomotor Retardation and Depression Severity at Follow-up (adjusting for baseline depression severity z-score).**

| **Study** | **Estimate** | **95% CI (Low)** | **95% CI (High)** | **P-value** | **n** |
| --- | --- | --- | --- | --- | --- |
| CADET | 0.145 | -0.022 | 0.311 | 0.09 | 527 |
| COBALT | 0.051 | -0.126 | 0.227 | 0.58 | 469 |
| GENPOD | -0.063 | -0.259 | 0.132 | 0.53 | 601 |
| IPCRESS | -0.362 | -0.628 | -0.096 | < 0.01 | 295 |
| MIR | -0.067 | -0.273 | 0.139 | 0.52 | 480 |
| PANDA | 0.174 | 0.030 | 0.319 | < 0.05 | 652 |
| REEACT | 0.127 | -0.028 | 0.281 | 0.11 | 685 |
| RESPOND | 0.224 | -0.076 | 0.523 | 0.15 | 220 |
| TREAD | -0.075 | -0.324 | 0.173 | 0.55 | 361 |

### **Table S4: Study-specific Regression Estimates for Psychomotor Retardation and Remission (adjusting for baseline depression severity z-score).**

| **Study** | **Odds Ratio** | **95% CI (Low)** | **95% CI (High)** | **P-value** | **n** |
| --- | --- | --- | --- | --- | --- |
| CADET | 0.712 | 0.480 | 1.167 | 0.13 | 527 |
| COBALT | 0.904 | 0.561 | 1.457 | 0.68 | 469 |
| GENPOD | 1.280 | 0.794 | 2.065 | 0.31 | 601 |
| IPCRESS | 1.550 | 0.754 | 3.186 | 0.23 | 295 |
| MIR | 1.210 | 0.727 | 1.999 | 0.47 | 480 |
| PANDA | 0.644 | 0.418 | 0.991 | < 0.05 | 652 |
| REEACT | 0.721 | 0.512 | 1.104 | 0.09 | 685 |
| RESPOND | 0.845 | 0.437 | 1.632 | 0.62 | 220 |
| TREAD | 1.070 | 0.591 | 1.928 | 0.83 | 361 |

**Figure S1: forest plot psychomotor retardation and primary outcome measure (adjusting for baseline depression severity z score).**

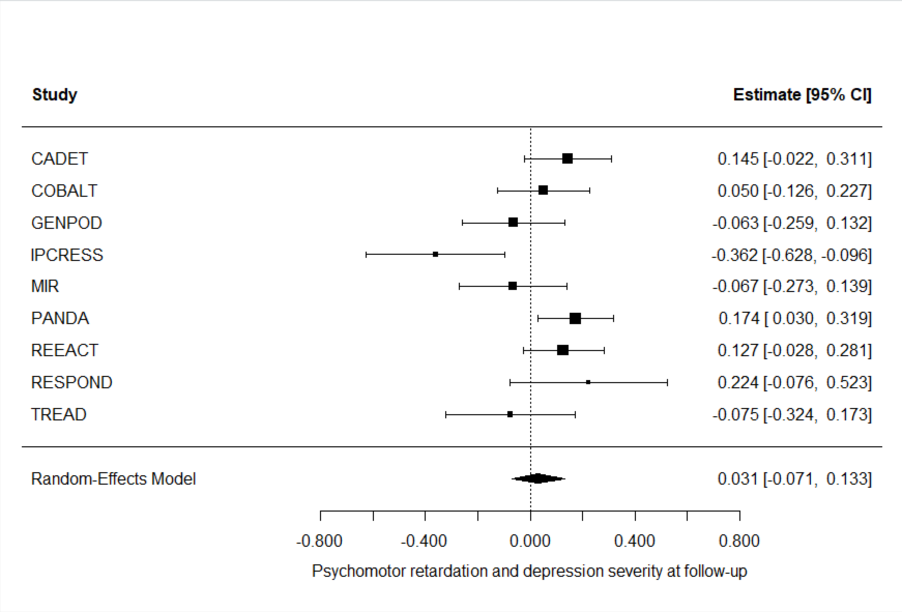
p = 0.552, I^2^ 59.1% (I^2^ 95% CI 9.4 – 91.5 %)

**Figure S2: forest plot psychomotor retardation and remission (adjusting for baseline depression severity z score).**

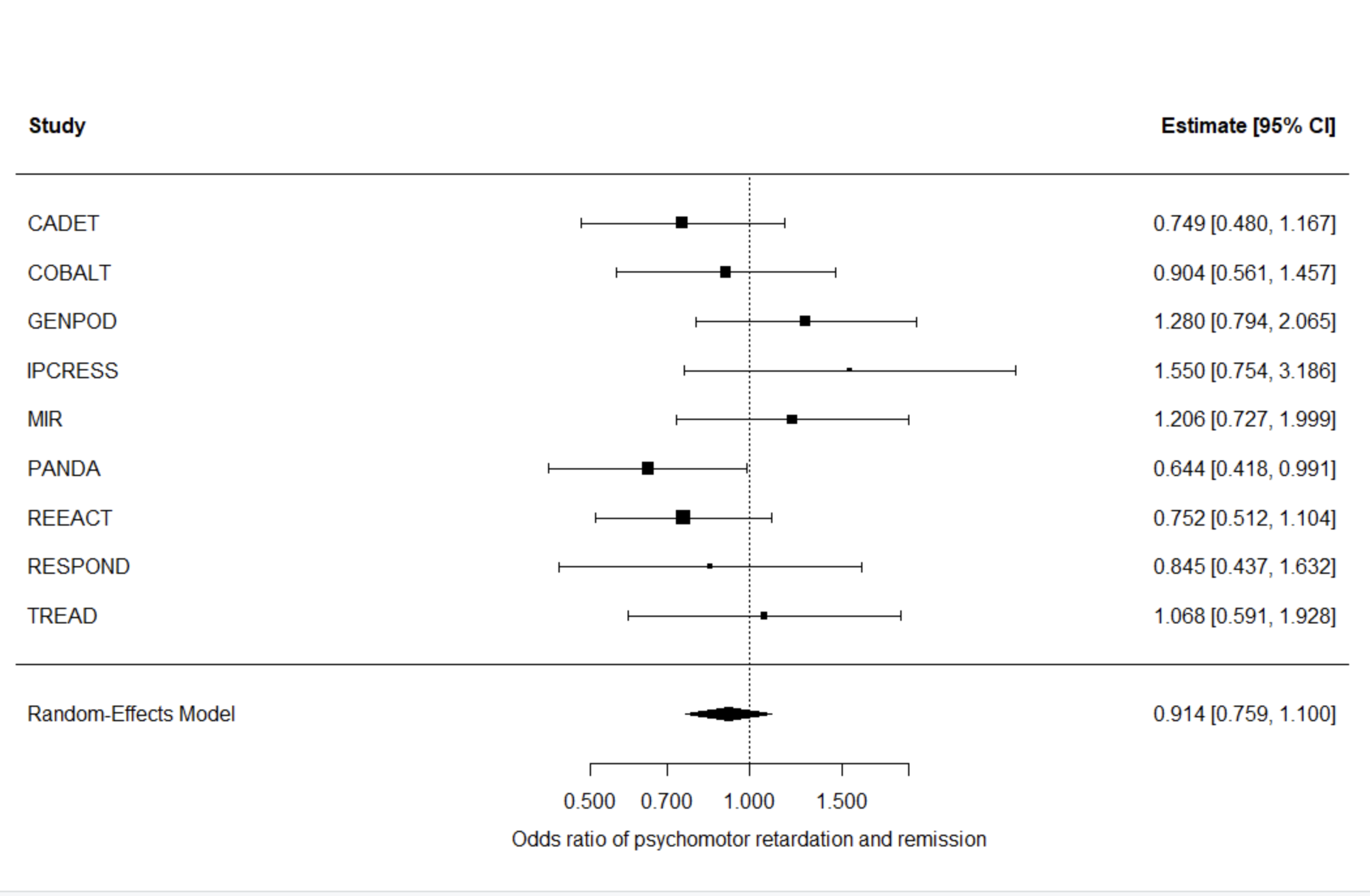

p = 0.342, I^2^ 19.5% (I^2^ 95% CI 0 – 78.3%)

**Figure S3: forest plot psychomotor retardation and primary outcome measure using imputed data (adjusting for CIS-R depression score).**

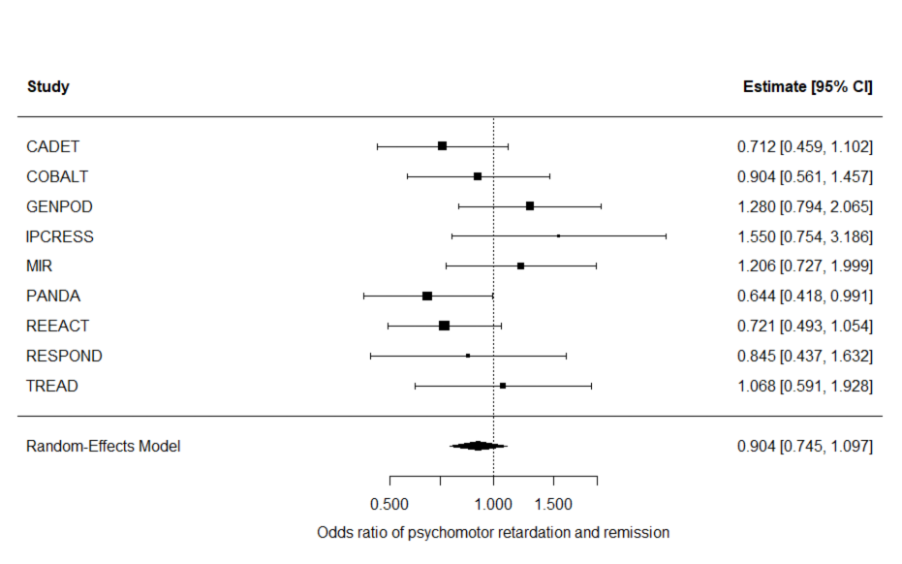

p = 0.308, I^2^ 26.2% (I^2^ 95% CI 0 – 80.1%)

**Figure S4: forest plot psychomotor retardation and remission using imputed data (adjusting for CIS-R depression score).**

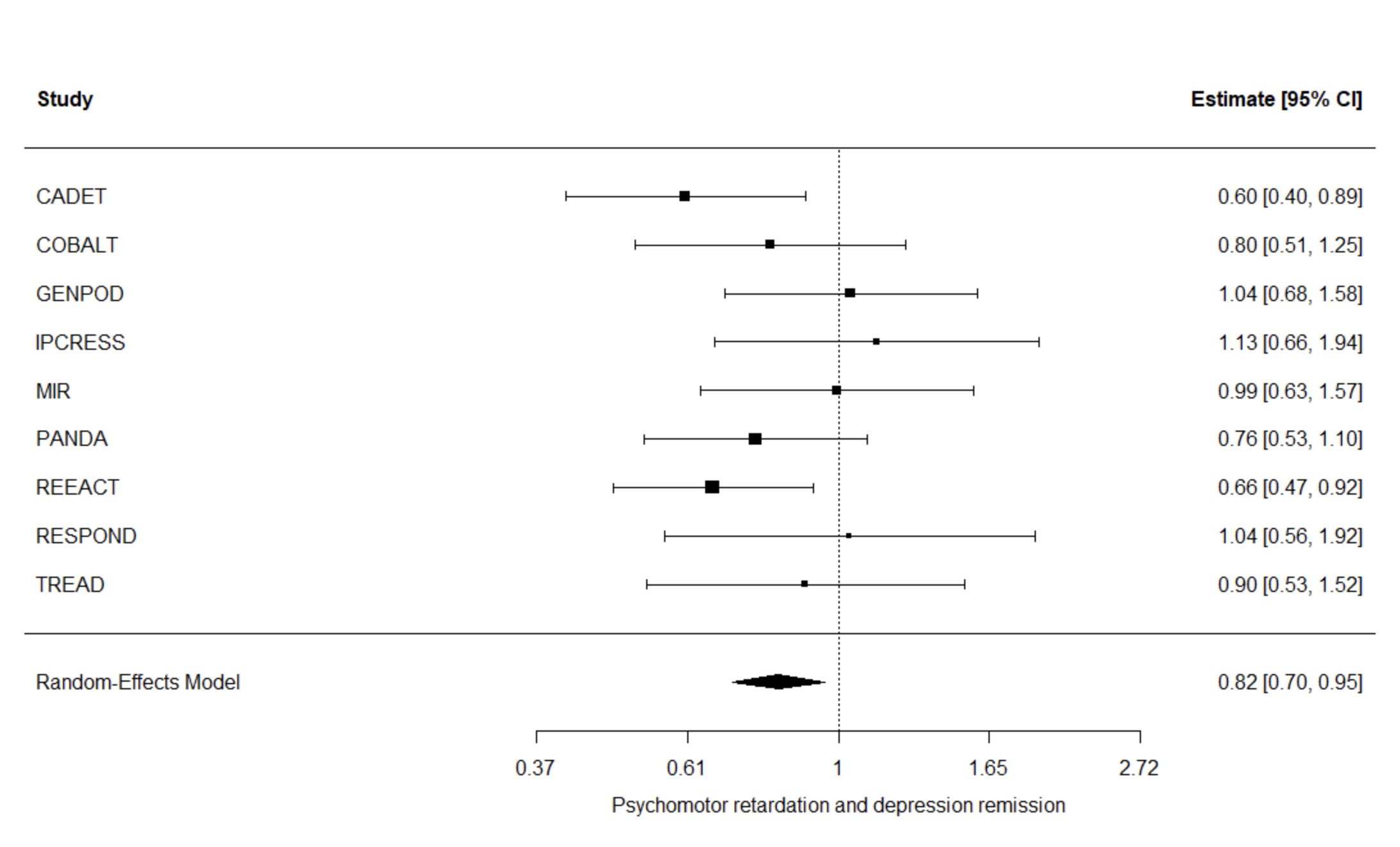

p < 0.05, I^2^ 9.78% (I^2^ 95% CI 0 – 72.1%)
